# Inhibitory KIR-Ligand Interactions and Relapse Protection Following HLA Matched Allogeneic HCT for AML

**DOI:** 10.1101/2020.07.10.20149047

**Authors:** Elizabeth Krieger, Rehan Qayyum, Amir Toor

## Abstract

Killer immunoglobulin-like receptor (KIR) and KIR-ligand (KIRL) interactions play an important role in natural killer cell mediated graft versus leukemia effect (GVL) after hematopoietic stem cell transplant (HCT) for AML. Mathematically accounting for KIR-KIRL interactions may identify donors with optimal NK cell mediated alloreactivity and GVL. In this retrospective study of 2359 donor recipient pairs (DRP) who underwent unrelated donor (URD) HCT for AML, KIR-KIRL interaction scores were determined. Relapse risk was significantly reduced in donor-recipient pairs (DRP) with higher inhibitory KIR-KIRL interaction and missing KIRL (mKIR) scores, with HR=0.86 (P=0.01) & HR=0.84 (P=0.02) respectively. This effect was not observed with activating KIR-KIRL interactions. The inhibitory KIR-KIRL (iKIR) interaction score components were summed to give an inhibitory-missing ligand (IM-KIR) score, which if it was 5 as opposed to <5, was also associated with a lower relapse risk, SHR 0.8 (P=0.004). Acute and chronic graft vs. host disease (GVHD), survival, GRFS and relapse free survival were not significantly different. However, TRM was increased among those with IM-KIR=5, HR, 1.32 (P=0.01). Among those with HLA matched DRP, anti-thymocyte globulin recipients with IM-KIR=5, had a lower relapse rate HR, 0.61 (p=0.001), however TRM was increased in these patients with a HR, 1.49 (p=0.034). This study demonstrates that unrelated DRPs with high inhibitory KIR content scores confer relapse protection, albeit with increased TRM. Clinical trials utilizing donors with a higher iKIR content in conjunction with novel strategies to reduce TRM should be considered for URD HCT recipients with AML to optimize clinical outcomes.

**Highlights:** Inhibitory and missing KIR-KIRL interactions, both individually and collectively are predictive of relapse following URD HCT for AML.

Amongst 8/8 HLA matched donor recipient pair relapse reduction was observed in those undergoing in vivo T cell depletion with ATG.

TRM is increased in patients with greater magnitude of inhibitory and missing KIR-KIRL interactions.

## Introduction

Acute myeloid leukemia (AML) is the most common form of adult acute leukemia and indication for hematopoietic cell transplantation (HCT) from human leukocyte antigen (HLA) matched unrelated donors (URD)^1,2^. Advances in donor selection, conditioning regimens and supportive measures have improved overall survival for patients with AML, however, relapse rates after HCT have remained stagnant^1,3–7,8^. The curative potential of HCT is predicated largely on graft vs leukemia (GVL) effect of the transplant^9^. Natural killer cells (NK) have been shown to provide cell-mediated alloreactivity, potentially by targeting non-antigen binding domains of the HLA molecules through germline-encoded receptors, such as, killer immunoglobulin-like receptors (KIR)^10,11^. These KIR have either inhibitory or activating function and interact with killer immunoglobulin-like receptor ligands (KIRL) on the target cells, specifically HLA molecules^12^. The effect of KIR-KIRL interactions in a HLA-matched URD HCT setting have long been debated with conflicting results ^13–22^. These inconsistencies may stem from the non-quantitative, descriptive methodology used to examine KIR-KIRL ligand interactions. These include a description of KIR-A and KIR-B haplotypes based on activating KIR gene content, and the presence of certain activating and inhibitory KIR genes and their ligands. Therefore, KIR genotyping, while commercially available, has not yet been established as a standard of care in unrelated donor selection as of yet, despite extensive laboratory evidence of the efficacy of NK cells in mediating anti-leukemia activity *in vitro*^23^.

This disconnect between known *in vitro* effective GVL mediation and apparent lack thereof *in vivo*, may be overcome by accounting for the complexity encountered in NK cell alloreactivity which is determined by multiple KIR and KIRL interacting simultaneously. A novel mathematical approach to quantifying KIR-KIRL interactions has been published^24^, utilizing a system of matrix, vector-operator equations to account for all possible donor-recipient KIR-KIRL interactions. This analysis highlighted the previously unknown importance of donor inhibitory KIR (iKIR) interactions, which complement the known missing KIRL interactions (mKIR) in mediating GVL.

In this study, the effect of known KIR-KIRL interactions on HCT outcomes in patients with AML is retrospectively examined in a large cohort of recipients of HLA matched and HLA mismatched URD to validate the findings reported earlier. Given the relevance of NK cells in mediating GVL effects following allogeneic HCT, the hypothesis that HCT for AML performed in donor-recipient pairs (DRP) with a higher donor inhibitory KIR-KIRL interaction would result in improved clinical outcomes, primarily by a reduction in relapse rates is examined.

## Methods

### Study design

The analysis presented here was performed on deidentified donor and recipient demographic and clinical outcomes data, provided by the Center for International Blood and Marrow Transplant Research (CIBMTR, Milwaukee, WI). In brief, data were available for 2,408 DRPs transplanted for AML between 2010 and 2016 at 113 centers across the United States. All donors were unrelated, and transplants facilitated via the National Marrow Donor Program. Patient variables examined included age, sex, Karnofsky performance status, HLA match grade (8/8 or 7/8, including HLA-A, -B, -C and -DRB1), KIR and HLA ligand status (A3, A11, Bw4 or Bw6, C1 or C2). KIR genotyping of all known KIR genes was available for all donors and was performed as previously described^25^. HLA typing was reported at high-resolution and KIR typing as presence or absence. Disease related characteristics included cytogenetic risk (SWOG criteria) and disease state at the time of transplant early (CR1) or late (CR 2-3). Cytogenetics were categorized as good, intermediate, poor; as a large number of observations were missing cytogenetics information (N=1494), a fourth category of ‘missing’ cytogenetics was included. Transplant-related variables included conditioning intensity, graft source, the use of in vivo T cell depletion (ATG or Alemtuzumab), and GVHD prophylaxis regimens utilized. Donor related characteristics included donor age, and KIR gene presence or absence.

### KIR KIRL interaction scores

KIR-KIRL interaction values were assigned as previously described^24,26^. Known KIR with established KIRL were utilized and are outlined in supplementary table 1. Essentially, the model assumes that NK cell interaction with its target cells may be mathematically described as a series of matrix equations, where the NK cell with its KIR molecules constitutes a ‘vector’, and the target with its KIRL molecules, an ‘operator’. The operator (target) modifies the vector (NK cell) upon interacting with it, either inhibiting or activating it. Each *individual* KIR-KIRL interaction may then be described as follows; if an inhibitory KIR (iKIR; represented by −1) on the NK cell encounters a ligand on its target (represented by 1), this will give the NK cell an inhibitory signal (overall effect −1). Mathematically this interaction which may then be represented by the equation, (−1) × (1) = −1, assuming a constitutively active basal state for NK cells. Conversely, if there is no ligand for an inhibitory KIR, i.e., missing KIRL (mKIRL; represented by −1), the interaction score will be (−1) × (−1) = +1 because of the abrogation of the inhibitory signal and persistent NK cell activation. Finally, activating KIR (aKIR; represented as +1) interacting with its ligands will be scored, (1) × (1) = +1, when the ligand is present, and (1) × (0) = 0, when the ligand is absent, since no activating signal is given. Each of these different scores constitutes a distinct component of the total KIR effect on individual NK cells expressing them, the cumulative effect determining the eventual outcome of NK cell – target cell interaction. The absolute value of each DRP’s KIR component scores, iKIR, mKIR and aKIR, were then summed to give the respective scores (**Supplementary Figure 1, Supplementary Table 2)**. The absolute value of each KIR component score was utilized to allow comparison of the relative magnitude of each component regardless of the direction of NK cell effect. This implies that iKIR score corresponds to the number of iKIR in the donor encountering their corresponding ligand in the recipient, while mKIR score is the number of iKIR in the donor missing their corresponding ligand in the recipient. The iKIR and mKIR scores were then summed up to calculate the composite inhibitory-missing ligand (IM)-KIR score, iKIR + mKIR = IM KIR. A logic-based algorithm (**Supplementary Table 3**) was utilized to calculate the KIR-KIRL interaction scores for the entire cohort in accordance with these equations and examples of iKIR, mKIR, aKIR and IM KIR Scores are given in Supplementary table 2.

### Outcome Definition

Relapse and relapse-free survival (RFS) were defined per CIBMTR criteria. Transplant related mortality (TRM) was defined as death in absence of relapse. Overall survival (OS) was defined as the time from HCT to last follow up or death from any cause. Chronic GVHD (cGVHD) was defined using NIH consensus criteria. GVHD free – relapse free survival was defined as freedom from relapse and cGVHD^27^.

### Statistical Methods

Characteristics between the patients who relapsed and who did not were compared using Pearson’s chi-square test or Student’s t-test. Kaplan-Meier survival curves by dichotomous IM-KIR scores were compared using the log-rank test. To evaluate the influence of confounding variables on the effect of NK cell mediated alloreactivity as determined by the KIR component scores, a multivariate Cox proportional hazards model was constructed including biologically relevant, clinical and demographic data available. Competing risk analysis treated TRM as a competing risk for relapse; analysis for TRM included relapse as a competing event; and for cGVHD occurrence, relapse and TRM before cGVHD diagnosis were treated analogously. Cox proportional hazard models were also generated using acute GVHD (grades 2-4 and 3-4), overall survival, and cGVHD-free, relapse-free survival as outcomes. These models were generated without and with adjustment for potential confounders based on prior literature and biological plausibility. To examine if the inclusion of missing cytogenetic data observations as a separate category might have affected the study results, sensitivity analyses were conducted using only observations without missing data. We performed additional exploratory analysis to understand the relationship between IM-KIR score and clinical outcomes in clinically important subgroups; these differences between the subgroups were examined using an interaction term in the models. All analyses were performed in Stata 15.0 (College Station, Texas) and a P-value <0.05 was considered significant. Since this study was designed to validate the findings reported previously, i.e., iKIR & mKIR individually and cumulatively impact clinical outcomes, multiple comparisons were not adjusted for.^31^

## Results

### Distribution of KIR-KIRL interaction scores in HLA matched URD SCT recipients

After excluding patients who failed to engraft neutrophils (N=30) or had missing data on relapse or time to relapse (N=12), the study cohort was comprised of 2365 donor recipient pairs (DRP) who underwent unrelated donor HCT for early or intermediate AML (**Table 1**). This cohort included adults aged 20-83 (mean 53) years; the majority (85%) of DRPs were high-resolution 8/8 HLA-matched for HLA-A, HLA-B, HLA-C, and HLA-DRB1. All patients received T cell replete grafts; 42% (n=996) received *in vivo* T cell depletion, 937 (94%) with ATG and 59 (6%) with Alemtuzumab. The majority 86% received a graft of mobilized peripheral blood stem cells (PBSC), 59% received myeloablative conditioning. KIR and KIRL gene frequencies within this population are given in **Supplementary Table 1**. This cohort was primarily of Caucasian descent (89%).

**Table 1:** Study population characteristics; total population and by IM-KIR scores.

| **VARIABLE** | **Study population§**<br>**(N=2,365)** | **IM-KIR<5**<br>**(N=1,418)** | **IM-KIR=5**<br>**(N=947)** | **P-value\*** |
| --- | --- | --- | --- | --- |
| **HLA 8/8 Match** | 2004 (84.7) | 1222 (86.2) | 782 (82.6) | 0.02 |
| **Cytogenetics** |  |  |  | 0.30 |
| *Good* | 142 (6.0) | 77 (5.4) | 65 (6.9) |  |
| *Intermediate* | 574 (24.3) | 334 (23.5) | 240 (25.3) |  |
| *Poor* | 155 (6.5) | 97 (6.8) | 58 (6.1) |  |
| *Missing* | 1494 (63.2) | 910 (64.2) | 584 (61.7) |  |
| **Disease Status** |  |  |  | 0.52 |
| *Early* | 1824 (77.1) | 1100 (77.6) | 724 (76.4) |  |
| *Intermediate* | 541 (22.9) | 318 (22.4) | 223 (23.5) |  |
| **Recipient Age** |  |  |  |  |
| 20-29 | 200 (8.5) | 118 (8.3) | 82 (8.7) | 0.74 |
| 30-39 | 254 (10.7) | 146 (10.3) | 108 (11.4) |  |
| 40-49 | 382 (16.1) | 236 (16.6) | 146 (15.4) |  |
| 50-59 | 600 (25.4) | 368 (25.9) | 232 (24.5) |  |
| ≥60 | 929 (39.3) | 550 (38.8) | 379 (40.0) |  |
| **CMV +Ve** | 1571 (66.9) | 943 (66.9) | 628 (66.9) | 0.98 |
| **KPS 90-100** | 1479 (63.4) | 864 (61.8) | 615 (65.8) | 0.04 |
| **Myeloablative Rx** | 1401 (59.2) | 853 (60.2) | 548 (57.9) | 0.27 |
| **Donor Age** |  |  |  |  |
| 18-19 | 93 (3.9) | 55 (3.9) | 38 (4.0) | 0.98 |
| 20-29 | 1268 (53.6) | 764 (53.9) | 504 (53.2) |  |
| 30-39 | 567 (24.0) | 338 (23.8) | 229 (24.2) |  |
| 40-49 | 295 (12.5) | 179 (12.6) | 116 (12.2) |  |
| ≥50 | 142 (6.0) | 82 (5.8) | 60 (6.3) |  |
| **In Vivo T-Cell Depletion** |  |  |  |  |
| **ATG** | 937 (39.6) | 561 (39.6) | 376 (39.7) | 0.99 |
| **Alemtuzumab** | 59 (2.5) | 35 (2.5) | 24 (2.5) |  |
| **None** | 1369 (57.9) | 822 (58.0) | 547 (57.8) |  |
| **GVHD Prophylaxis** |  |  |  |  |
| **Tacrolimus-Based** | 2091 (88.4) | 1239 (87.4) | 852 (90.0) | 0.05 |
| **Cyclosporine-Based** | 274 (11.6) | 179 (12.6) | 95 (10.0) |  |
| **Graft Source** |  |  |  | 0.19 |
| **Bone Marrow** | 342 (14.5) | 216 (15.2) | 126 (13.3) |  |
| **PBMC** | 2023 (85.5) | 1202 (84.8) | 821 (86.7) |  |
| **Sex Match** |  |  |  | 0.01 |
| **Male to Male** | 929 (39.3) | 577 (40.7) | 352 (37.2) |  |
| **Male to Female** | 708 (29.9) | 438 (30.9) | 270 (28.5) |  |
| **Female to Male** | 298 (12.6) | 157 (11.1) | 141 (14.9) |  |
| **Female to Female** | 430 (18.2) | 246 (17.4) | 184 (19.4) |  |
| **Relapse** | 947 (40.0) | 481 (33.9) | 266 (28.1) | 0.003 |
| **Death** | 1175 (49.7) | 711 (50.1) | 464 (49.0) | 0.59 |

\*- P values for differences between IM-KIR-2,3,4 and IM-KIR5 populations.
§- Of the 361, who got a 7/8 HLA matched donor HCT, 163 (45%) were mismatched for HLA-A, 82 (23%) for HLA-B, 63 (17%) for HLA-C and 53 (15%) for HLA-DRB1.

Absolute values of the KIR-KIRL interaction scores varied across the population regardless of the HLA match grade (8/8 vs 7/8) and different ethnic groups of donors and patients. Absolute values of the KIR-KIRL interaction component scores were; iKIR median 3 (range: 1 to 5); mKIR median 2 (range: 0 to 4); aKIR median 0 (range: 0 to 4). The median IM-KIR score was 4, (range: 2 to 5) (**Figure 1**).

**Figure 1.**
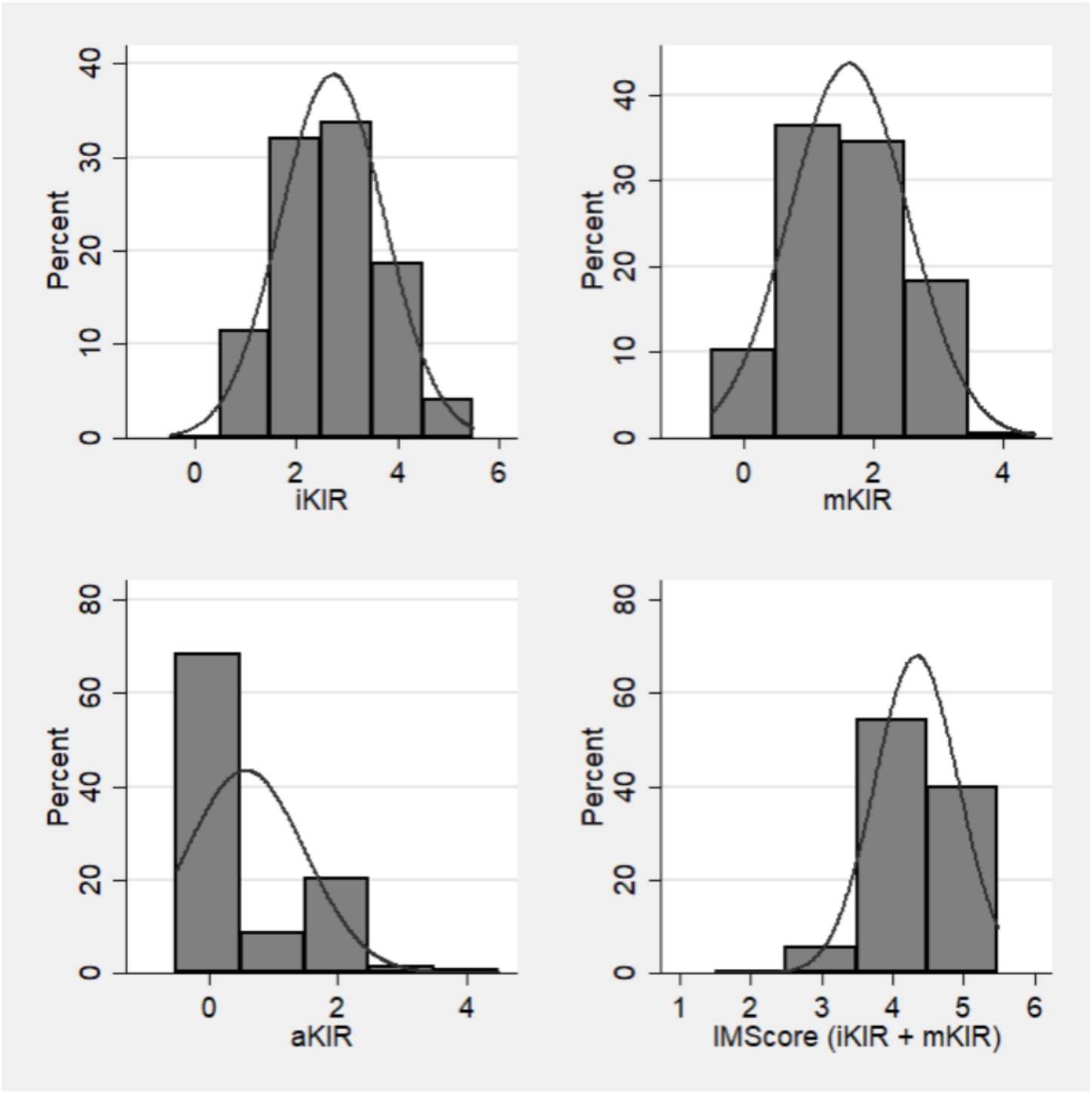
Population frequencies of KIR-KIRL interaction scores for inhibitory KIR (iKIR), missing KIRL (mKIR), activating KIR (aKIR) and IM-KIR scores. (N=2316).

### Inhibitory KIR scores impact relapse risk following HLA matched URD SCT

The effect of all 3 KIR-KIRL interaction component scores (absolute values of iKIR, mKIR and aKIR) on relapse was evaluated as a continuous variable. Higher iKIR and mKIR scores, were significantly associated with a reduced risk of AML relapse, with sub-hazard ratios (SHR) of 0.86 and 0.85 respectively (**Figure 2**). However, aKIR did not significantly impact relapse risk for AML. These effects were observed independent of several confounding variables included in the multivariable analysis, specifically, recipient and donor ages, conditioning regimen intensity and *in vivo* T cell depletion, which if unfavorable (older donor age, RIC, and ATG use) all increased the risk for relapse. No significant effect of the KIR-KIRL interactions was observed on other clinical outcomes examined (acute GVHD, overall and relapse-free survival or TRM). The likelihood of cGVHD was higher in those with a higher iKIR score (SHR 1.09), albeit not statistically significantly.

**Figure 2.**
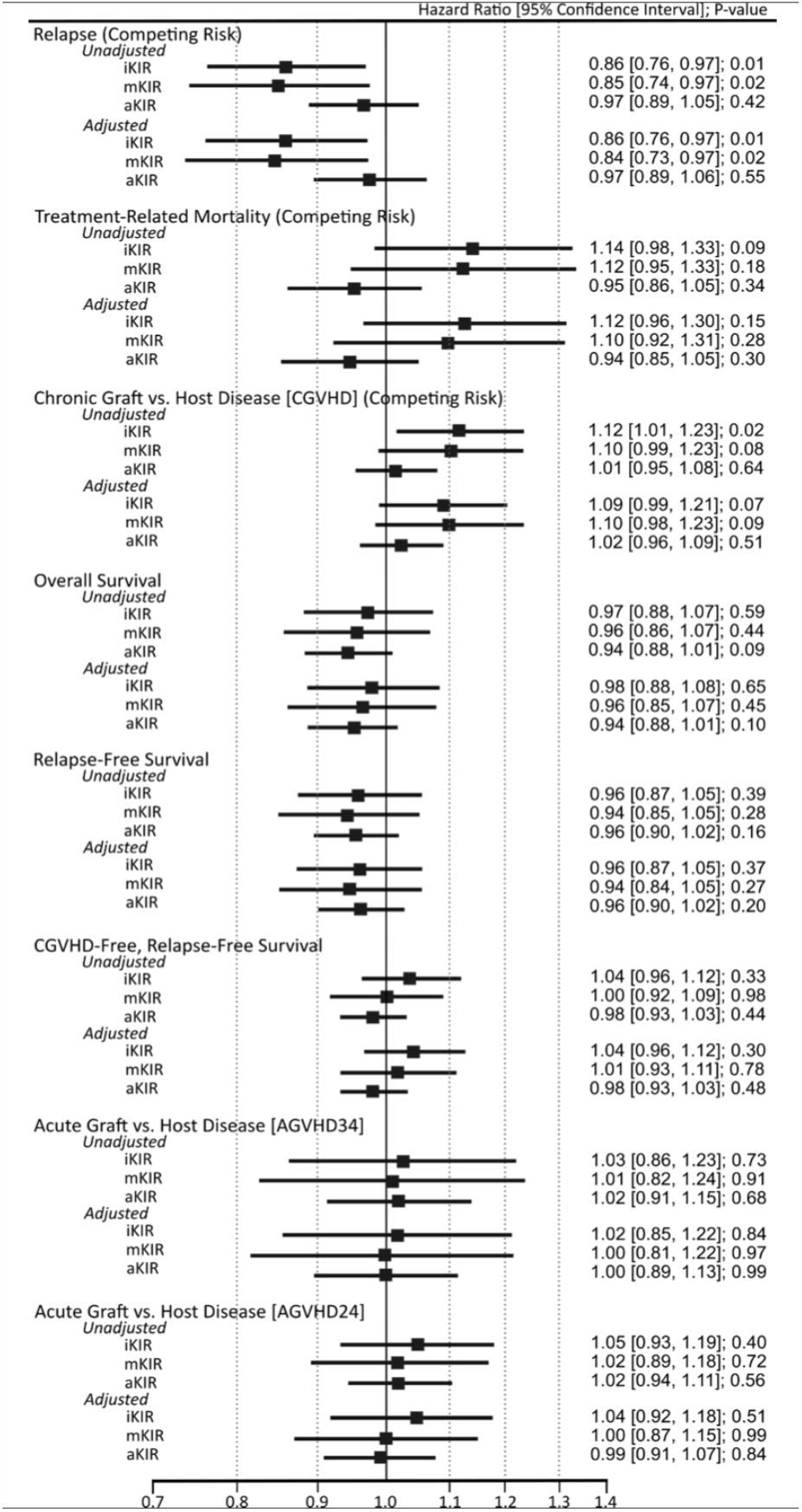
Unadjusted and adjusted sub-distribution hazard and hazard ratios from competing risk and Cox proportional hazard models for the effect of iKIR, mKIR and aKIR on clinical outcomes following HCT for AML. Sensitivity analysis performed to account for missing values of confounding variables.

### A dichotomous IM-KIR score delineates relapse risk

Given their independent predictive value for relapse following URD HCT, the cumulative effect of iKIR and mKIR component scores was then examined with the IM-KIR score. IM KIR score was dichotomized for analysis into those DRP with an IM KIR score of 5 (IM=5), and those with an IM-KIR scores of 2, 3 or 4 (IM<5). The cut-off value was chosen to have relatively balanced groups as few DRPs had IM KIR scores of 2 or 3 (**Figure 1**). As compared with DRPs in the IM<5 group, patients in the IM=5 group had a 20% lower risk of relapse (SHR 0.80**; Figures 3 & 4**). Patients in the IM=5 group remained at a lower risk of relapse, (SHR 0.80), when adjusted for other relevant demographic variables (**Table 2**). As observed with the two component scores (iKIR & mKIR), the relapse protection was independent of older recipient and donor ages (>49 years), ATG use and RIC usage, which were all associated with increased relapse risk.

**Table 2:**
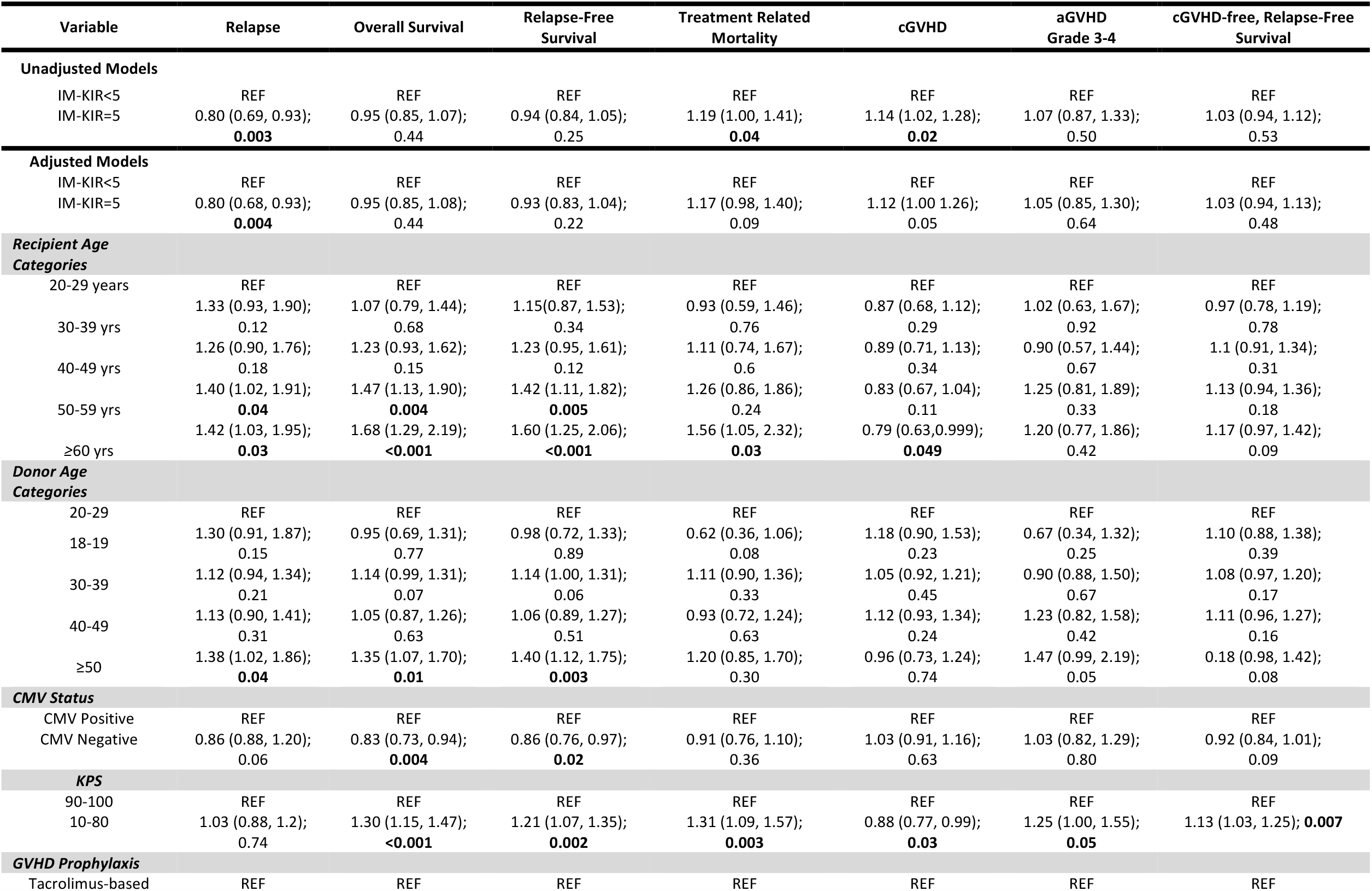
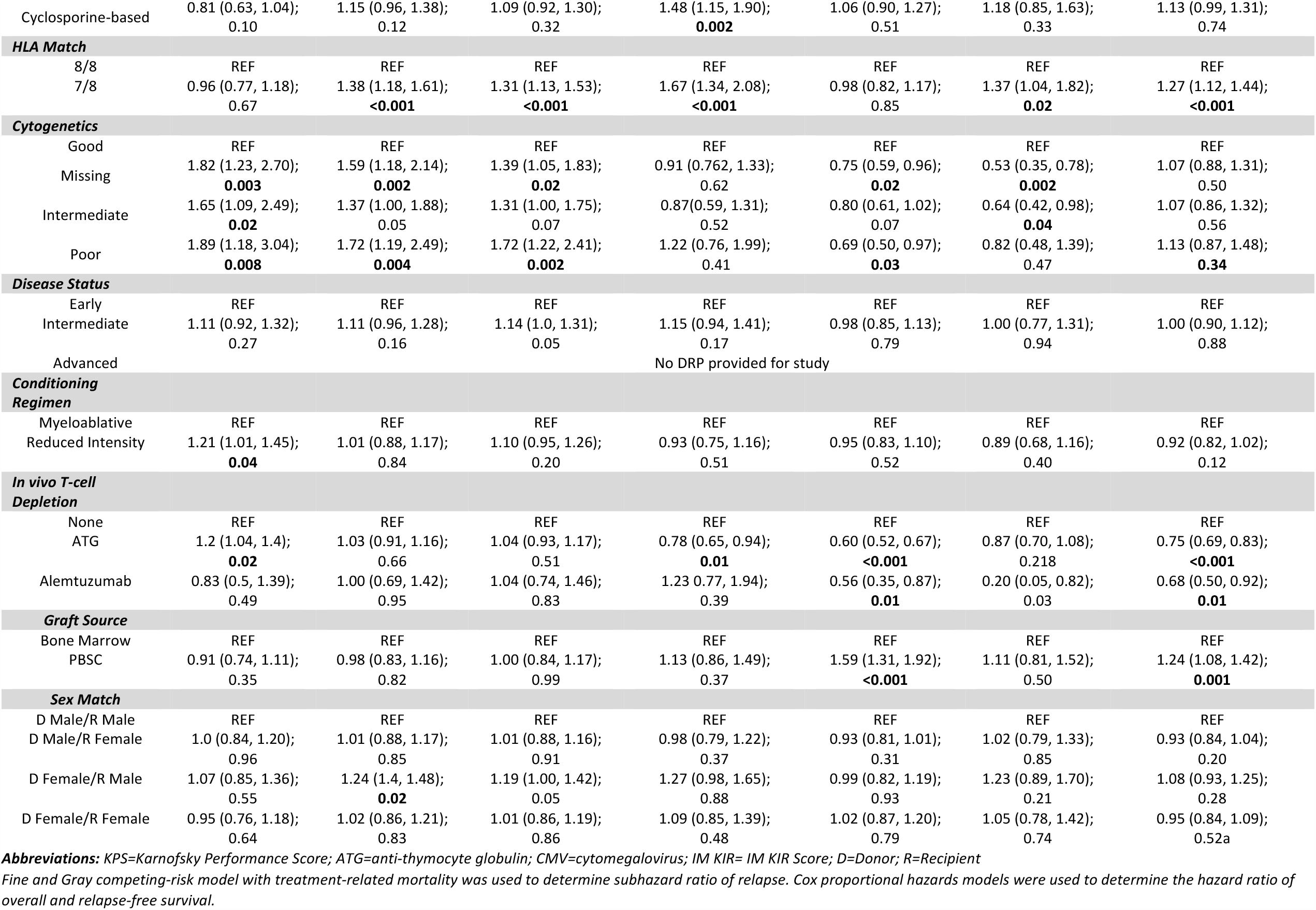
Unadjusted and adjusted results of the competing risk and Cox proportional hazard models for the effect of IM-KIR score on clinical outcomes for URD HCT for AML. SHR (95% CI); P-values reported. During the 1,965,798 person-day follow up, 747 patients relapsed, and 1,175 patients died.

**Figure 3.**
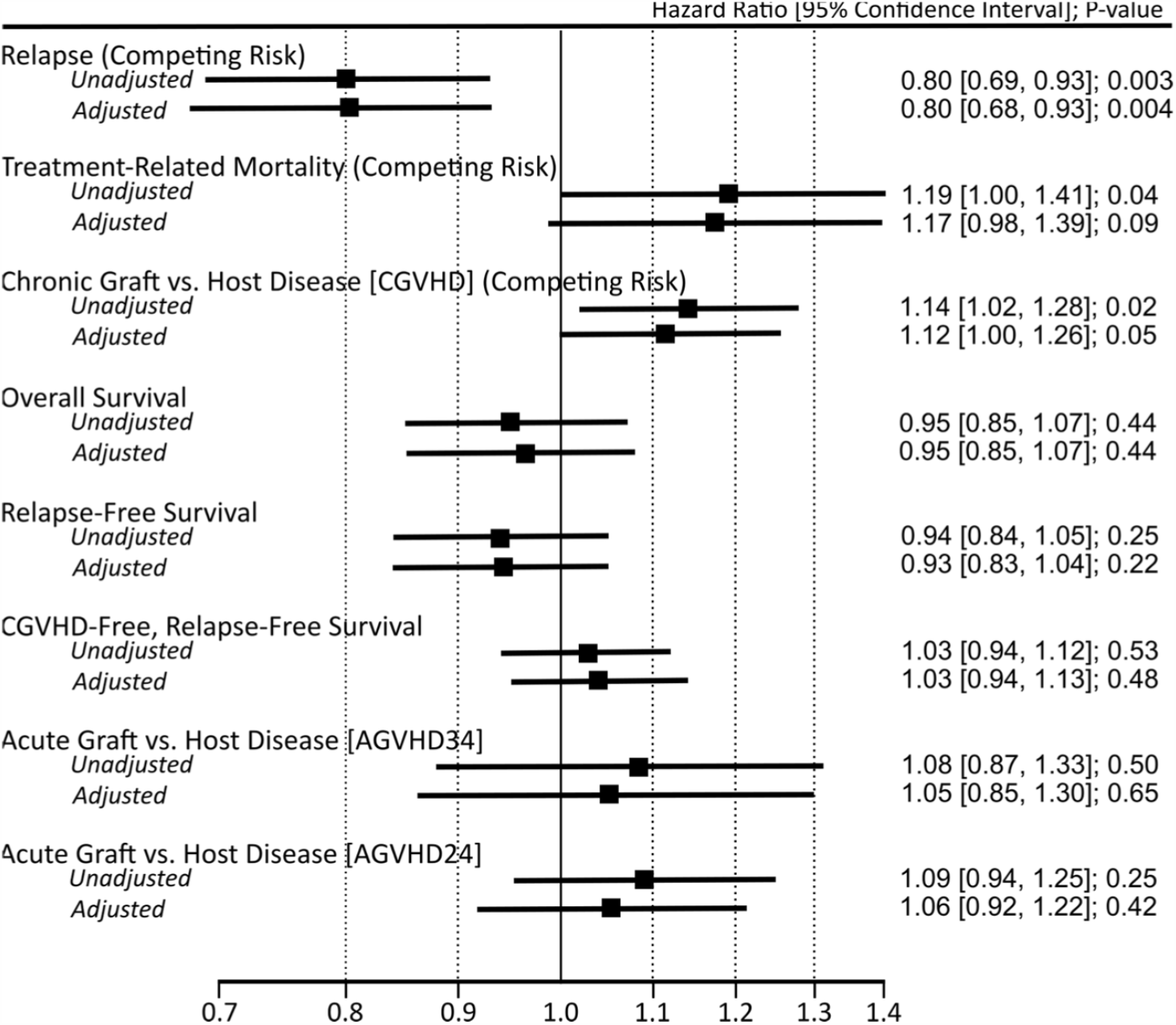
Unadjusted and adjusted sub-distribution hazard and hazard ratios from competing risk and Cox proportional hazard models for the effect of DIM Scores on clinical outcomes following HCT for AML. Sensitivity analysis performed to account for missing values of confounding variables.

**Figure 4.**
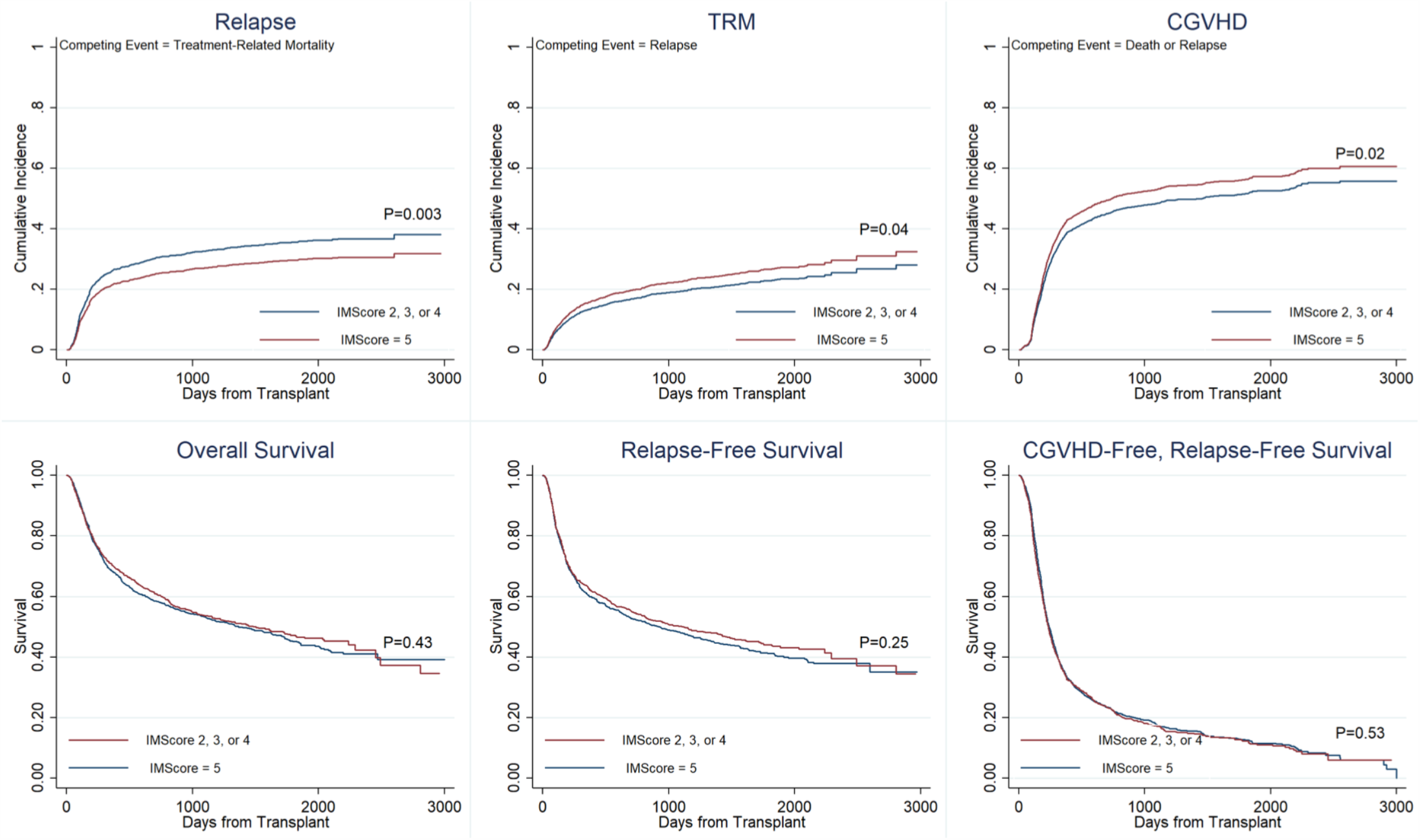
Unadjusted cumulative incidence curves by IM-KIR scores for relapse, treatment-related mortality, and chronic graft versus host disease, as well as failure curves by IM-KIR scores for overall survival, relapse-free survival, and cGVHD-free, relapse-free survival.

Patients in the IM=5 group also had a higher risk of developing cGVHD (SHR 1.13) in unadjusted analysis (**Figure 3 & 4**), but, as with individual KIR-KIRL interaction components this did not remain statistically significant on multivariate analysis (**Table 2**). Acute GVHD was also not impacted by the IM-KIR score. TRM risk was increased in the IM=5 patients, but this difference was not independent of increased recipient age, poor KPS, HLA mismatch and cyclosporine-based GVHD prophylaxis which, increased the risk for TRM (**Table 2**). Despite the strong relapse protection effect, there was not a survival or relapse-free survival benefit observed within the two IM-KIR score categories.

### IM KIR score and relapse protection in recipients of HLA matched allografts

To determine the effect of variation in IM-KIR score and outcomes in distinct groups of patients, interaction analyses were performed. First, the interaction between IM-KIR score and degree of HLA matching was examined in the entire cohort. In those transplanted with an 8/8 HLA matched donor, who were also IM=5, there was a significant reduction in relapse risk (SHR 0.76, p=0.001) as compared to the reference group (IM<5), however this was not so among the 7/8 HLA match and IM=5 donor recipients (**Supplementary Figure 2**). The interaction analysis did not disclose a significant relationship between the degree of HLA mismatch and relapse reduction in the IM=5 patients (interaction P =0.12). As the degree of HLA match did not modify IM-KIR effect on relapse protection, and in general HLA mismatch was associated with poor survival (**Table 2**), further subset analysis was limited to 8/8 HLA-matched (n=1958) DRP.

Among those transplanted with an 8/8 HLA matched donor, 730 patients received ATG for *in vivo* T cell depletion: of those, the patients who received grafts from IM<5 donors, 38% had relapsed within 3 years, while among those transplanted with IM=5 donors, only 26% relapsed (SHR 0.61) (**Table 3, Figure 5**). Notably, the IM-KIR score for the donor modified the effect of the use of ATG on relapse risk (interaction p=0.049), i.e., the DRP with an IM=5 donor, when administered ATG, did not experience the increased relapse risk seen in IM<5 DRP who received ATG (**Table 3, Figure 5**). On the other hand, among those patients who did not receive any *in vivo* T cell depletion the 3-year relapse rates were not significantly different in those transplanted with IM=5 (27%) vs. IM<5 (29%) donors. Relapse reduction was also seen in IM=5 DRP among most subgroups of patients examined (**Table 3**).

**Table 3:** Subgroup analysis of 8/8 HLA matched DRP, with alemtuzumab patients removed. Unadjusted and adjusted results of the competing risk, Cox proportional hazard models and interaction analysis for the effect of IM-KIR score on clinical outcomes for URD HCT for AML. SHR (95% CI); P-values reported.

| Variable | Relapse | Overall Survival | Relapse-Free Survival | Treatment Related Mortality | cGVHD | aGVHD Grade 3-4 | cGVHD-free, Relapse-Free Survival |
| --- | --- | --- | --- | --- | --- | --- | --- |
| **Unadjusted Models** |  |  |  |  |  |  |  |
| IM-KIR=5 | **0.75 (0.64, 0.89); 0.001** | 0.96 (0.85, 1.1); 0.61 | 0.93 (0.82, 1.06); 0.27 | **1.29 (1.01, 1.58); 0.01** | **1.14 (1.01, 1.29); 0.04** | 1.17 (0.92, 1.48); 0.19 | 1.04 (0.95, 1.15); 0.39 |
| **Adjusted Models** |  |  |  |  |  |  |  |
| IM-KIR=5 | **0.74 (0.63, 0.89); 0.001** | 0.99 (0.87, 1.13); 0.91 | 0.94 (0.83, 1.07); 0.35 | **1.32 (1.09, 1.62); 0.006** | 1.12 (0.98, 1.26); 0.11 | 1.15 (0.91, 1.46); 0.24 | 1.05 (0.95, 1.17); 0.31 |
| **Subgroup Analyses - To determine the effect of Subgroups on the relationship between IM-KIR and outcomes** |  |  |  |  |  |  |  |
| **In vivo T cell Depletion** |  |  |  |  |  |  |  |
| IM-KIR=5 |  |  |  |  |  |  |  |
| ATG (730) | **0.61 (0.46, 0.81); 0.001** | 0.90 (0.72, 1.13); 0.37 | 0.84 (0.68, 1.04); 0.11 | **1.49 (1.03, 2.16); 0.034** | 1.00 (0.79, 1.27); 0.98 | 1.07 (0.72, 1.6); 0.73 | 0.97 (0.82, 1.16); 0.77 |
| None (1188) | 0.87 (0.7, 1.08); 0.21 | 1.04 (0.88, 1.23); 0.63 | 1.01 (0.86, 1.19); 0.89 | 1.25 (0.97, 1.59); 0.08 | 1.15 (1.0, 1.34); 0.07 | 1.15 (0.85, 1.56); 0.36 | 1.12 (0.98, 1.27); 0.09 |
| Interaction P value | **0.049** | 0.3 | 0.15 | 0.31 | 0.42 | 0.82 | 0.19 |
| **Conditioning Intensity** |  |  |  |  |  |  |  |
| IM-KIR=5 |  |  |  |  |  |  |  |
| Myeloablative (1131) | **0.77 (0.61, 0.97); 0.03** | 1.04 (0.87, 1.26); 0.65 | 1.02 (0.86, 1.22); 0.79 | **1.48 (1.12, 1.96); 0.006** | 1.10 (0.94, 1.3); 0.24 | **1.4 (1.02, 1.91); 0.04** | 1.12 (0.98, 1.29); 0.08 |
| Reduced intensity (787) | **0.71 (0.55, 0.91); 0.01** | 0.92 (0.76, 1.13); 0.44 | 0.84 (0.69, 1.01); 0.07 | 1.15 (0.85, 1.56); 0.35 | 1.12 (0.92, 1.36); 0.27 | 0.91 (0.62, 1.33); 0.62 | 0.96 (0.82, 1.22); 0.62 |
| Interaction P value | 0.63 | 0.42 | 0.14 | 0.24 | 0.95 | 0.1 | 0.11 |
| **Graft Source** |  |  |  |  |  |  |  |
| IM-KIR=5 |  |  |  |  |  |  |  |
| Bone Marrow (274) | 0.87 (0.52, 1.46); 0.6 | 1.11 (0.75, 1.65); 0.59 | 1.13 (0.77, 1.65); 0.54 | 1.66 (0.86, 3.2); 0.13 | 1.43 (0.94, 2.13); 0.1 | 1.28 (0.57, 2.88); 0.54 | **1.40 (1.03, 1.9); 0.03** |
| PBSC (1644) | **0.72 (0.6, 0.87); 0.001** | 0.96 (0.83, 1.11); 0.59 | 0.9 (0.78, 1.03); 0.13 | **1.27 (1.03, 1.58); 0.03** | 1.09 (0.95, 1.24); 0.21 | 1.14 (0.88, 1.47); 0.31 | 1.01 (0.91, 1.13); 0.83 |
| Interaction P value | 0.31 | 0.23 | 0.075 | 0.25 | 0.37 | 0.8 | **0.03** |
| **Disease state at the time of transplant** |  |  |  |  |  |  |  |
| IM-KIR=5 |  |  |  |  |  |  |  |
| Early | **0.78 (0.64, 0.95); 0.012** | 1.01 (0.87, 1.18); 0.83 | 0.96 (0.83, 1.11); 0.59 | **1.29 (1.02, 1.62); 0.03** | 1.11 (0.96, 1.28); 0.15 | 1.17 (0.89, 1.53); 0.27 | 1.07 (0.95, 1.2); 0.27 |
| Intermediate | **0.67 (0.45, 0.99); 0.049** | 0.98 (0.72, 1.33); 0.89 | 0.997 (0.75, 1.33); 0.98 | **1.71 (1.09, 2.68); 0.02** | 1.08 (0.81, 1.45); 0.58 | 1.34 (0.79, 2.28); 0.28 | 1.02 (0.81, 1.29); 0.84 |
| Interaction P value | 0.41 | 0.62 | 0.74 | 0.44 | 0.91 | 0.91 | 0.99 |

Relative adjustments made for recipient age, donor Age, CMV status, KPS score, GVHD prophylaxis regimen, cytogenetics, disease state at the time of transplant, conditioning regimen, In vivo T cell depletion, graft source and sex match.

**Figure 5.**
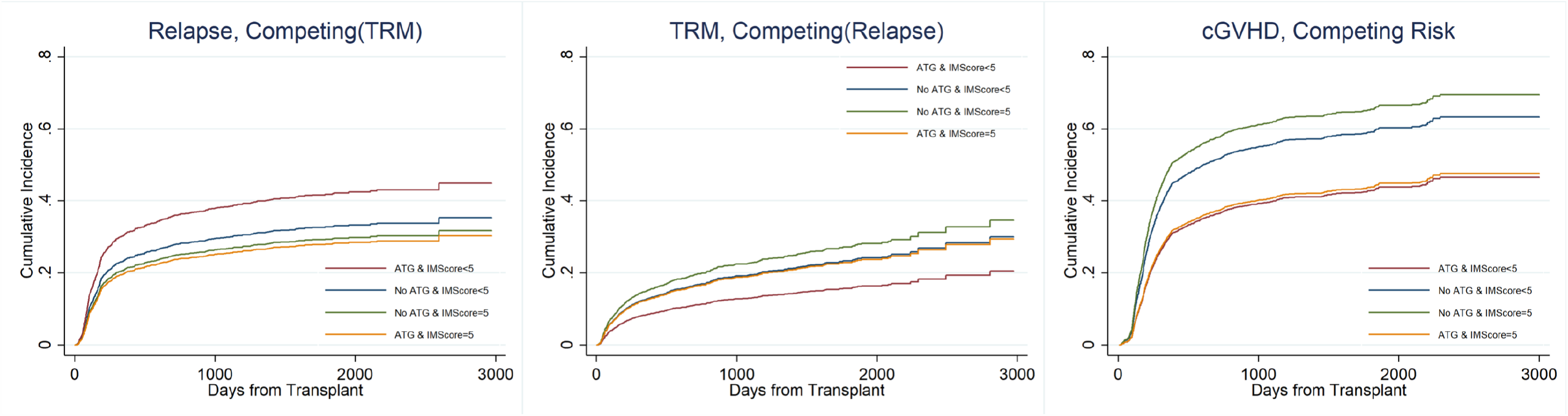
Unadjusted cumulative incidence curves by IM-KIR scores and in vivo T cell depletion with ATG depicting relapse, treatment-related mortality, and chronic graft versus host disease.

### IM-KIR score influence on transplant related mortality and GVHD

Despite the relapse risk reduction seen in IM=5 donors, there was an absence of survival, RFS and GRFS benefit. This is related to the increased TRM observed in the 8/8 HLA matched, IM=5 DRP as compared to IM<5 donors (SHR 1.32; **Table 3**). This effect appeared to be most prominent among those who received ATG and had IM=5 donors (SHR 1.49; **Table 3; Figure 5)**. The TRM increase may be contributed to by a higher, albeit non-significant likelihood of cGVHD developing in those who did not receive ATG and had an IM=5 donor as opposed to IM<5 (SHR 1.15; **Table 3, Figure 5**).

## Discussion

In this study a novel, logic-based mathematical-model examining the KIR-KIRL interactions is reported as a surrogate measure of NK cell alloreactivity in patients with AML undergoing URD HCT. In this large cohort of HLA matched and single antigen mismatched DRPs, a higher iKIR and mKIR score, as well as combined IM-KIR score predicted a lower risk of relapse in recipients transplanted from a donor with a higher iKIR content. This relapse risk reduction was most evident in HLA matched donor-recipient pairs (DRP) undergoing transplantation with *in vivo* T cell depletion using ATG. Despite this relapse risk reduction, no survival advantage was observed, due to a higher risk of TRM observed in the DRP with the highest inhibitory KIR content.

The mathematical approach to examining KIR-KIRL interactions utilized here is similar to the model system utilized in studying the interaction of T cell receptors with minor histocompatibility antigen-HLA complexes^28^. The vector (NK cell) – operator (target cell) interaction model allows for all the known interactions to be quantified in a sound mathematical framework, and the clinical associations of this cumulative interaction studied. Somewhat contrary to the current dogma, this analytic methodology has identified a strong association of iKIR donor-recipient interactions with AML relapse protection. It is known that *in vitro* models have shown that NK cells with an increased number of iKIRs that interact with their requisite HLA are more responsive when they encounter a target cell ^29,30^. This suggests that cells reconstituting in a post HCT setting with the potential for a larger number of iKIR-HLA interactions may potentially have higher graft vs leukemia (GVL) activity. The results reported here support that counterintuitive notion, which may be understood using the analogy of an electrical capacitor. In this instance the iKIR interactions of a NK cell may be considered analogous to the dielectric of a capacitor. Like a capacitor which stores electric charge, NK cells may be considered to possess alloreactive killing potential. Capacitors with a stronger dielectric (insulator) capacity can store a larger amount of charge, likewise NK cells with increased iKIR interactions, in the early post-transplant setting may be inhibited by the vast majority of cells with appropriate iKIRL on their surfaces and retain a larger ‘killing capacity’ for when they encounter a potential target. Downregulation of HLA, class I and II molecules on AML blasts is well described immune evasion mechanism for escape from T cell alloreactivity^31–33^. However, the *primed* iKIR-rich-NK cells upon encountering such blasts may provide more robust effector function.

Further, in the converse situation NK cells with a greater aKIR content which encounter aKIRL will experience constant activating input (weaker dielectric and insulation), and potentially be activated in a nonspecific fashion, eventually being tuned down and rendered anergic to prevent auto-reactivity^34^. This quantitative perspective of NK cell interactions with normal vs. leukemic cells would suggest that iKIR interactions may increase the specificity of the NK cell alloreactivity, making it more likely to effect a GVL response as observed here (**Figure 6**). Indeed, the results reported here suggest that the greater the iKIR content of the donor, the greater the benefit in terms of relapse protection, regardless of the number of iKIR ligands present.

**Figure 6.**
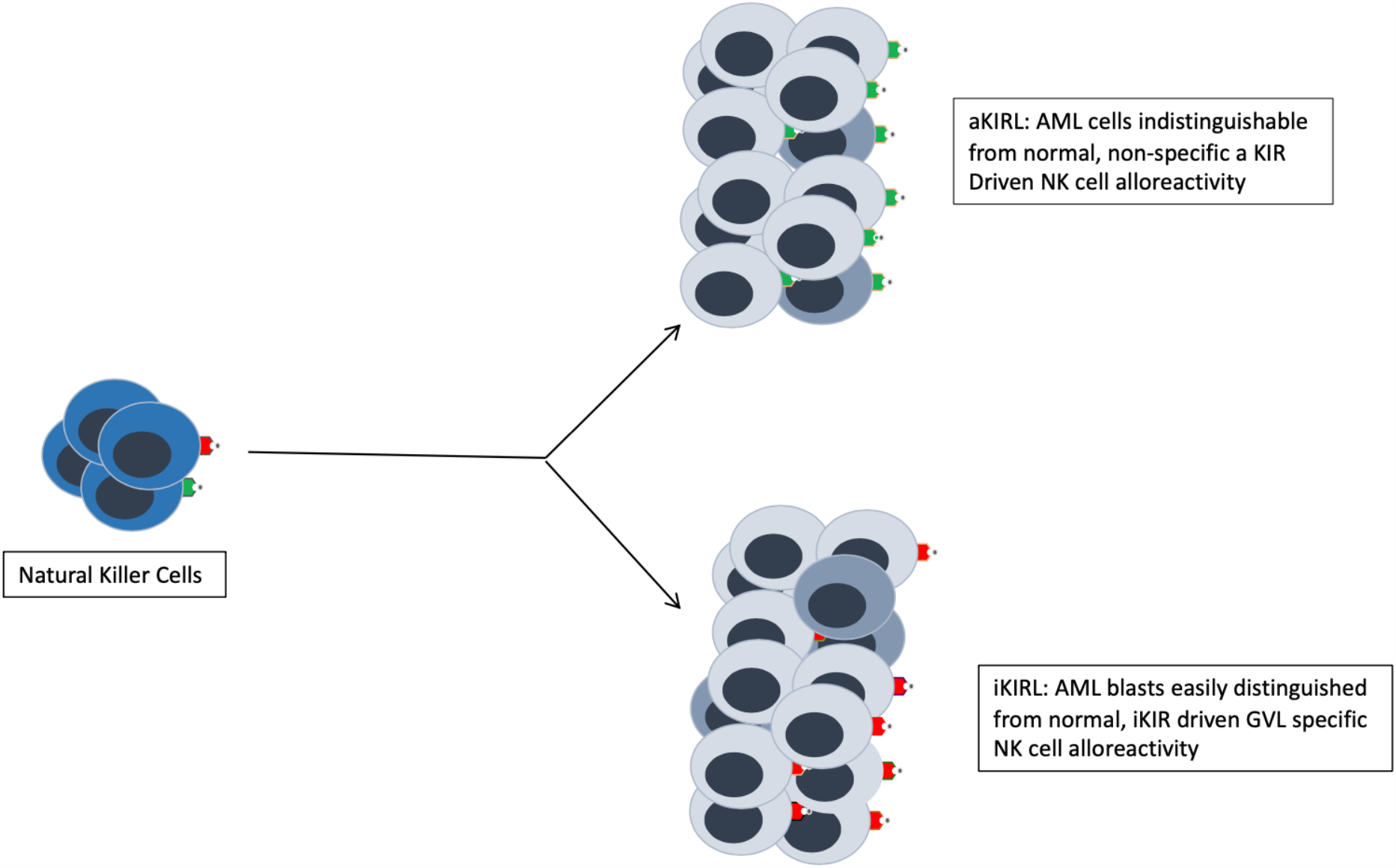
NK cell alloreactivity model. Numerous activating aKIR-KIRL interactions (green receptors & ligands) between aKIR expressing NK cells (Blue) and normal (light grey) as well as malignant cells (dark grey), imply that the NK cells cannot specifically target leukemic blasts, which may lead to non-specific and thus, ineffective NK cell mediated alloreactivity. With iKIR-KIRL interactions (red receptors & ligands) where activation only occurs upon interaction with iKIRL ^low/negative^ cells, more specific and targeted GVL activity may be observed.

The relapse benefit of the IM=5 was most evident and significant in HLA matched allograft recipients of *in vivo* T cell depleted (TCD) grafts where ATG was used. This was in contrast with the overall higher relapse risk in ATG recipients in the cohort as a whole, and in IM<5 donors in particular. Further, the IM=5 and IM<5 patients who did not get ATG had no significant difference in relapse risk, albeit at a higher risk of developing cGVHD. One may reason that, higher NK cell alloreactivity of the greater iKIR and mKIR interactions (IM=5) attenuated the higher relapse risk observed when ATG was given and cGVHD mediated alloreactivity and relapse protection therefrom neutralized. Similar relapse protection was observed in recipients of RIC conditioning, suggesting an NK cell mediated alloreactive mechanism of relapse reduction.

Despite the lower relapse risk no survival benefit of IM=5 donors observed in this study, rather, a higher TRM was seen. The cumulative incidence of TRM in recipients given ATG with IM=5 donors is higher than in those getting ATG with an IM<5 donor, who had the lowest TRM risk, despite an equal rate of cGVHD with the ATG and IM=5 donors. This suggests that the additional TRM risk may be related to infections in donors with higher iKIR content, rather than GVHD. From an evolutionary standpoint, NK cells protect the host from viral infections^35–38^. Many viruses, in particular the Herpesviridae, such as cytomegalovirus and Epstein-Barr virus have developed elaborate mechanisms for evading NK cell and CD8+ T cell mediated lysis of infected cells. These include deployment of decoy HLA class I receptors, such as UL 18 in CMV which would increase the inhibitory input to the NK cells. IM=5 as opposed to IM<5 donor derived NK cells may be more susceptible to such viral suppression of NK cell effect of combating viral infection, contributing to TRM. Interestingly, amongst HLA matched patients those who received PBSC, and had donors with IM=5, a lower relapse rate was observed, without increases in TRM, perhaps because of the higher T cell dose delivered with these grafts.

HLA matching has long been the gold standard for URD selection, with an ever increasing level of precision in matching achieved through high resolution typing^39^. However beyond that there has been little which distinguishes equivalently HLA matched donors, until recently when large data sets established donor age to be of definite prognostic value in URD transplant outcomes^40,41^. Even though it is well established that NK cells reconstitute early following HCT^42^, and the effect of KIR-KIRL interactions on NK cell mediated alloreactivity has been well established^16,22,43^, KIR genotyping has not been successfully applied to donor selection^44^. Greater activating KIR content or KIR B haplotypes have been reported to have a favorable influence on clinical outcomes^45,46^. However, in this large cohort, iKIR and mKIRL scores, had a protective role *vis a vis* relapse. The data presented here give a mathematical framework for incorporating KIR genotyping in URD selection for future clinical trials.

This model is simple, and its underlying mathematics provide the ability to grow and adapt as new discoveries are made regarding NK cell alloreactivity and KIR interactions in a post-transplant setting, particularly for the aKIR for which the ligands are not known yet. However, it is important to note that at this moment it considers all KIR HLA interactions uniformly. It is known that some KIR, for example, KIR 2DL2 binds HLA C with a higher affinity then KIR 2DL3, this means that the model may underestimate or overestimate the effect of specific interactions^47,48^. The different relative contributions of certain KIR-KIRL interactions will need to be examined in future studies to overcome this limitation but can easily be accommodated in the mathematical framework of the model utilized. Effects of NK cell receptors like NKG2 family of receptors and their ligands on the target cells may also be incorporated in this vector – operator model system. Further, these data also raise an important mechanistic questions, why should iKIR and mKIR have the same magnitude of effect? Aspects of NK cell biology not accounted for in this model may contribute to this; for example, NK cell education, or, the time dependent expression of KIR following HCT, as well as any effect KIRL-zygosity might have on these interactions.

In conclusion, a strong relapse protection effect was observed in patients transplanted with donors with an IM-KIR score of 5. This challenges the notion that KIR are irrelevant to donor selection, and raises the question that those donors with the highest IM KIR scores in an appropriate transplant setting, may be considered as optimal donors for HCT recipients with AML to allow for increased NK cell mediated alloreactivity. Future clinical trials evaluating donor selection for URD HCT should include this measure to evaluate its value prospectively in uniformly treated patient cohorts, with adequate GVHD and antiviral prophylaxis to mitigate TRM.

## Data Availability

The data are available through the CIBMTR.

## Acknowledgements

The authors gratefully acknowledge Stephen Spellman and Tao Wang, PhD for their help with data organization and for critical review of the manuscript. The analysis presented here was performed on data obtained from the Center for International Blood and Marrow Transplantation Research (CIBMTR, Milwaukee, WI). The CIBMTR is supported primarily by Public Health Service grant/cooperative agreement U24CA076518 with the National Cancer Institute (NCI), the National Heart, Lung and Blood Institute (NHLBI) and the National Institute of Allergy and Infectious Diseases (NIAID); grant/cooperative agreement U24HL138660 with NHLBI and NCI; grant U24CA233032 from the NCI; grants OT3HL147741, R21HL140314 and U01HL128568 from the NHLBI; contract HHSH250201700006C with Health Resources and Services Administration (HRSA); grants N00014-18-1-2888, N00014-17-1-2850 and N00014-20-1-2705 from the Office of Naval Research; subaward from prime contract award SC1MC31881-01-00 with HRSA; subawards from prime grant awards R01HL131731 and R01HL126589 from NHLBI; subaward from prime grant award 5R01CA2151343 from NIH Cancer Institute. P01CA111412, R01CA152108, R01CA218285;R01CA231141, R01HL126589, R01HL129472, R01HL131731 and U01AI126612 from the NIH. The views expressed in this article do not reflect the official policy or position of the National Institute of Health, the Department of the Navy, the Department of Defense, Health Resources and Services Administration (HRSA) or any other agency of the U.S. Government. AT was supported by research funding from the NIH-NCI Cancer Center Support Grant (P30-CA016059; PI: Gordon Ginder, MD). Institutional Review Board at the Virginia Commonwealth University deemed that since data were completely deidentified, this analysis did not constitute human subjects research and according to Section 45 CFR 46.102(l) of the HHS Regulations for the Protection of Human Subjects did not require IRB scrutiny.

EK: Designed study, developed KIR-KIRL scoring algorithm, analyzed data, wrote paper.

RQ: Designed study, developed KIR-KIRL scoring algorithm, performed statistical analysis, wrote paper.

AT: Designed study, developed KIR-KIRL scoring algorithm, analyzed data, wrote paper.

---

**Supplementary Table 1.** KIR and corresponding KIRL population frequency in the study cohort.

| **KIR AND CORRESPONDING KIRL FREQUENCIES IN STUDY POPULATION\*** |  |  |  |
| --- | --- | --- | --- |
| **KIR** | **Frequency (%)** | **KIRL** | **Frequency (%)** |
| **2DL1** | 97 | **C2** | 85 |
| **2DL2** | 52 | **C1** | 60 |
| **2DL3** | 90 | **C1** | 60 |
| **3DL1** | 95 | **BW4** | 60 |
| **3DL2** | 100 | **HLA A3/A11** | 26%/10% |
| **2DS1** | 39 | **C2** | 85 |
| **2DS2** | 53 | **HLA A11** | 10 |
| **2SD4** | 95 | **HLA A11** | 10 |
| **2DS5** | 31 | **C2** | 85 |

**Supplementary Table 2:** Examples of KIR scoring in 2 potential DRP.

| *Donor iKIR* | *Recipient HLA* | *Score* | *iKIR* | *mKIR* | *aKIR* | *Absolute value* |
| --- | --- | --- | --- | --- | --- | --- |
| 2DL1(-1) | C2 (+1) | -1 | -1 |  |  | 1 |
| 2DL2(-1) | Absent (-1) | +1 |  | +1 |  | 1 |
| 2DL3(-1) | Absent (-1) | +1 |  | +1 |  | 1 |
| 3DL1(-1) | Absent (-1) | +1 |  | +1 |  | 1 |
| 3DL2(-1) | Absent (-1) | +1 |  | +1 |  | 1 |
|  |  |  |  |  |  | IM KIR<br>Score=5 |
| 2DS1(+1) | C2(+1) | +1 |  |  | +1 |  |
| 2DS2(+1) | Absent (0) | 0 |  |  |  |  |
| 2DS4(+1) | Absent (0) | 0 |  |  |  |  |
| 2DS5(+1) | C2(+1) | +1 |  |  | +1 |  |
|  |  |  | *iKIR=-1* | *mKIR=+4* | *aKIR=+2* |  |
| 2DL1(-1) | C2(+1) | -1 | -1 |  |  | 1 |
| - | C1(+1) | 0 |  |  |  |  |
| 2DL3(-1) | C1(+1) | -1 | -1 |  |  | 1 |
| 3DL1(-1) | Bw4(+1) | -1 | -1 |  |  | 1 |
| 3DL2(-1) | A3(+1) | -1 | -1 |  |  | 1 |
|  |  |  |  |  |  | IM KIR<br>Score=4 |
| 2DS1(+1) | C2(+1) | +1 |  |  | +1 |  |
| 2DS2(+1) | -(0) | 0 |  |  |  |  |
| -(0) | -(0) | 0 |  |  |  |  |
| -(0) | C2(+1) | 0 |  |  |  |  |
|  |  |  | *iKIR=-4* | *mKIR=0* | *aKIR=+1* |  |

**Supplementary Table 3.**
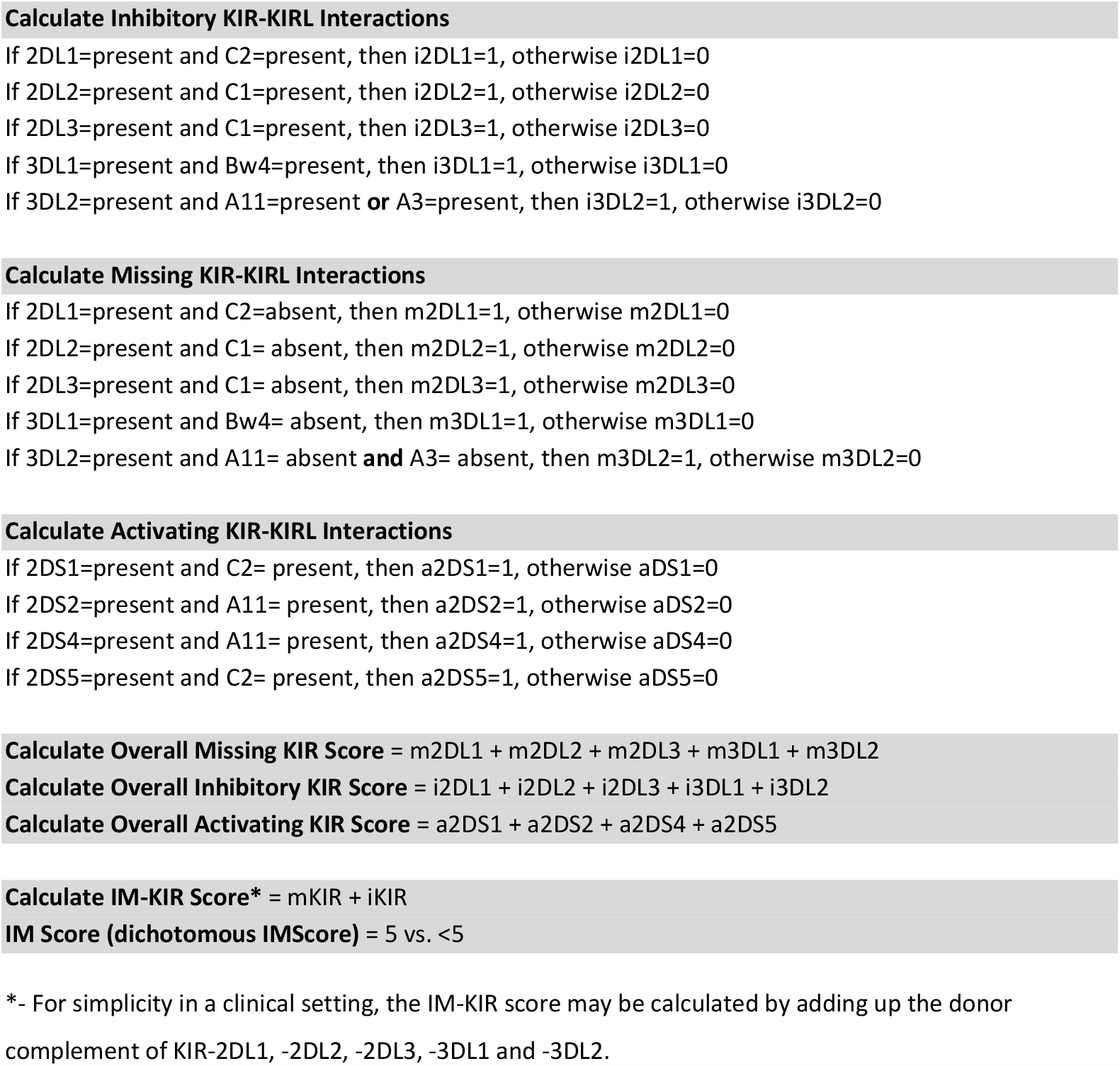
Algorithm to generate KIR-KIRL interaction component scores and the IM-KIR score.

**Supplementary Figure 1.**
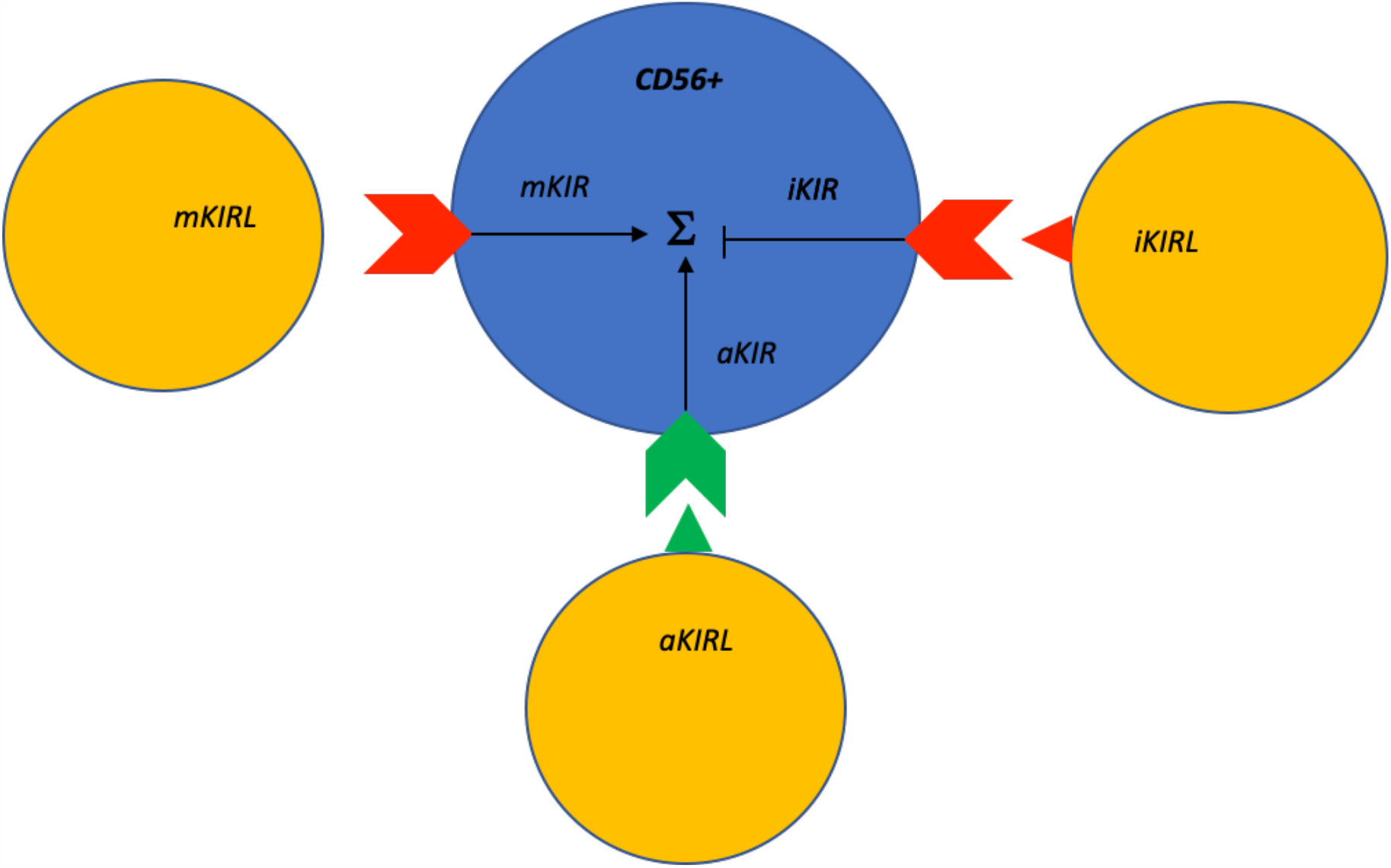
NK cell (blue) with inhibitory KIR (red chevron) and activating KIR (green chevron) interacting with targets which either express, or do not express cognate ligands (red and green triangles). The KIR input collectively determines NK cell function, i.e., NK cell mediated alloreactivity.

**Supplementary Figure 2.**
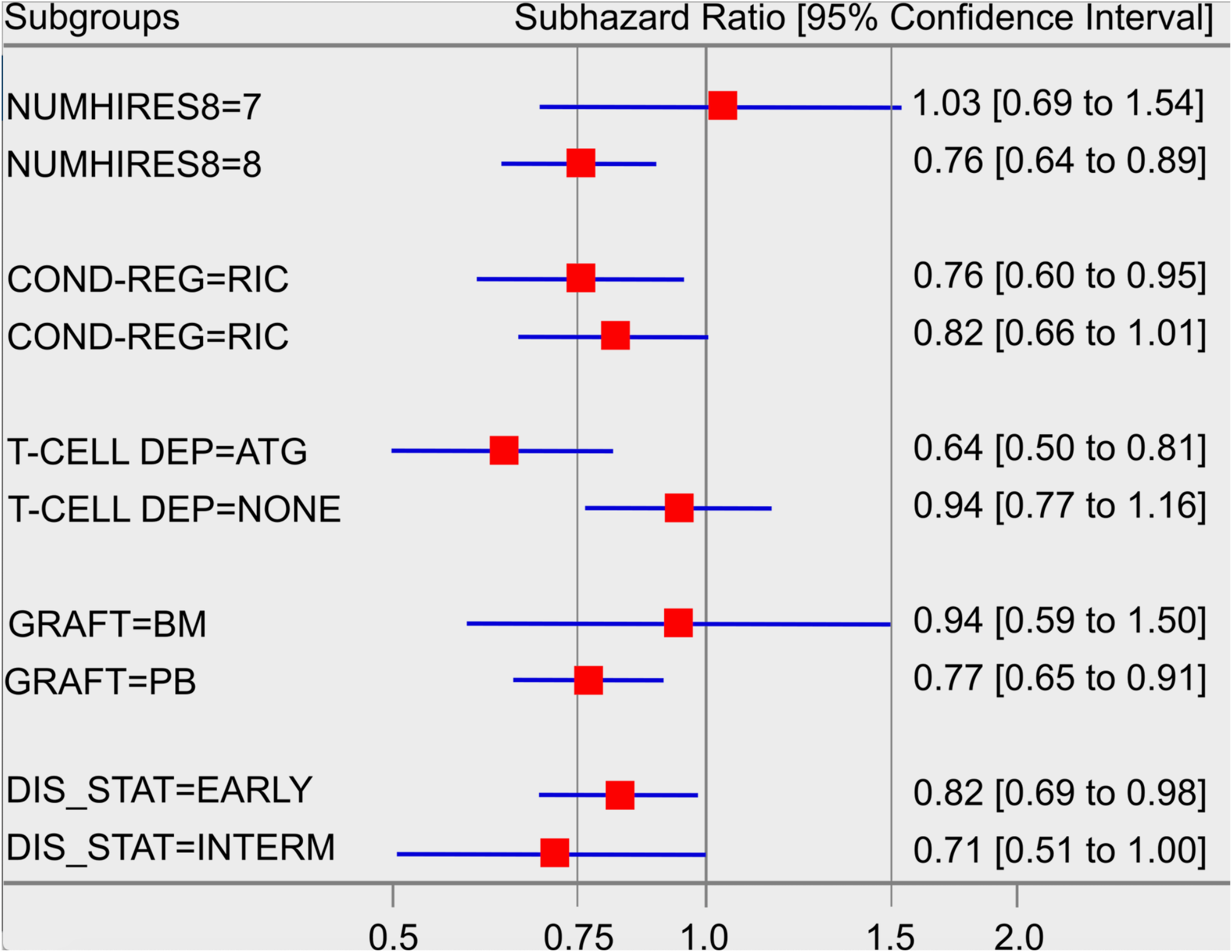
Adjusted sub-distribution hazard and hazard ratios for relapse from competing risk and Cox proportional hazard models for the effect of IM=5 on clinical outcomes following HCT for AML in subgroups.

